# Valproate-induced hyperammonemic encephalopathy versus asymptomatic hyperammonemia in adults with epilepsy: a cross-sectional study

**DOI:** 10.64898/2026.09.05.26362327

**Authors:** Huijuan Chen, Xuan Wang

**Author notes:** **Corresponding author:** Xuan Wang, Department of Neurology, Affiliated Hospital of Jiangsu University, Zhenjiang, Jiangsu Province, 212000, China.

## Abstract

**Background and Objective:** To compare clinical features of valproate (VPA)-induced hyperammonemic encephalopathy and asymptomatic hyperammonemia to inform monitoring strategies.

**Methods:** This cross-sectional study at Affiliated Hospital of Jiangsu University included 25 adults with epilepsy hospitalized between January 2024 and March 2026 with serum ammonia >33 μmol/L during VPA therapy. Nine met predefined VPA-induced hyperammonemic encephalopathy (VHE) criteria; 16 had asymptomatic hyperammonemia. Clinical, laboratory, electroencephalography (EEG), and neuroimaging parameters were compared.

**Results:** VHE patients had higher ammonia (median 76.0 [IQR 51.2–155.7] vs 42.5 [33.5–66.0] μmol/L; P<0.001) and shorter VPA duration (median 0 [0–60] vs 30 [0–120] months; P=0.005) than asymptomatic patients. Crucially, VPA trough concentrations and liver enzymes did not differ between groups. Diffuse EEG slowing occurred exclusively in VHE patients(66.7% vs 0%; P<0.001); prior cerebral infarction was more frequent in VHE (88.9% vs 43.8%; P=0.040). All VHE patients improved clinically within 1–3 days and achieved ammonia normalization within 3–10 days after VPA withdrawal without specific ammonia-lowering therapy.

**Conclusions:** VHE typically emerges early in VPA therapy, is unpredictable by routine therapeutic drug monitoring or liver function tests, and resolves rapidly upon discontinuation. Clinicians should maintain a low threshold for ammonia testing in patients with new neurological symptoms during early VPA treatment, particularly those with prior cerebrovascular disease or diffuse EEG slowing. These findings support integrating ammonia screening into VPA safety protocols beyond standard therapeutic drug monitoring.

**Key Points:**

1. Elevated blood ammonia levels during valproate therapy do not reliably predict the presence of brain-related symptoms, as some patients remain entirely asymptomatic despite significant laboratory abnormalities.
2. Clinical decisions should be guided by observable changes in mental status and behavior rather than ammonia test results alone, to avoid both unnecessary treatment and delayed recognition of true drug-induced encephalopathy.
3. Routine monitoring for adults on valproate should pair laboratory testing with regular assessment of cognitive function, ensuring that symptomatic cases are promptly identified while avoiding over-intervention in stable patients.

## 1. Introduction

Epilepsy is one of the most common chronic neurological disorders worldwide[1]. In China, more than 9 million people are affected, with approximately 650,000–700,000 new cases diagnosed annually (incidence 50–70 per 100,000 and prevalence approximately 5 per 1,000)[2–4], making it the second most common neurological disease after cerebrovascular disease[5, 6]. Most patients require long-term treatment with anti-seizure medications (ASMs), among which valproic acid (VPA) is widely used as a first-line agent for both generalized and focal epilepsies owing to its broad-spectrum efficacy, favorable tolerability, and low cost[7, 8]. With the expanding use of VPA, its adverse effects—particularly hyperammonemia and valproate-induced hyperammonemic encephalopathy (VHE)—have drawn increasing clinical attention.

VHE is a rare but potentially serious complication of VPA therapy, typically presenting with acute or subacute altered mental status and neuropsychiatric symptoms accompanied by elevated serum ammonia, often in the absence of overt liver dysfunction [9, 10]. The proposed pathophysiologic mechanisms include inhibition of carbamoyl phosphate synthetase I, a rate-limiting enzyme of the hepatic urea cycle, and enhanced renal tubular glutaminase activity, resulting in increased ammonia production and reduced ammonia elimination [11]. However, the existing literature has focused predominantly on case reports and small case series of patients with overt encephalopathy, leaving the entity of “asymptomatic hyperammonemia”—that is, elevated serum ammonia without clinical features of encephalopathy—largely undercharacterized [12]. Retrospective and cross-sectional studies have reported hyperammonemia in a substantial proportion (ranging from 16% to over 40%) of VPA-treated patients, yet the majority remain asymptomatic, and their clinical profiles are poorly defined [13, 14]. Consequently, clinicians may overlook this condition and misattribute patients’ behavioral abnormalities to inadequately controlled seizures or comorbid psychiatric disorders, thereby delaying recognition and timely intervention [15].

In this exploratory study, we characterized the clinical profile of patients with epilepsy who developed hyperammonemia during VPA therapy and compared those with and without encephalopathy across demographic characteristics, VPA treatment history, laboratory parameters, electroencephalography (EEG), and neuroimaging findings. We hypothesized that (1) asymptomatic hyperammonemia is not uncommon among VPA-treated patients, (2) VHE occurs independently of VPA trough concentrations and liver enzyme elevations, and (3) diffuse EEG slowing is associated with symptomatic presentation. Given the paucity of comparative data on asymptomatic hyperammonemia, this study was designed as a hypothesis-generating analysis to inform risk stratification and ammonia monitoring strategies in VPA-treated patients.

## 2. Methods

### 2.1. Study design and ethics

This was an exploratory, single-center, cross-sectional study of consecutive adult inpatients with epilepsy, conducted in the Department of Neurology, Affiliated Hospital of Jiangsu University, between January 2024 and March 2026. Owing to the exploratory nature of the study and the rarity of VHE, no a priori sample size calculation was performed; all eligible patients during the study period were enrolled.

The study protocol was reviewed and approved by the Institutional Ethics Committee of the Affiliated Hospital of Jiangsu University (approval number: KY2024K1102) and was performed in accordance with the ethical standards of the 1964 Declaration of Helsinki and its later amendments. Written informed consent was obtained from all participants or their legal guardians before enrollment. All patient data were de-identified prior to analysis. As this was a purely observational study without investigator-controlled intervention, registration in a public clinical trial registry was not required.

### 2.2. Study population

Consecutive patients with epilepsy who were hospitalized in the Department of Neurology, were receiving VPA, and had a serum ammonia concentration exceeding the institutional upper limit of normal (>33 μmol/L) were enrolled. Eligible Patients were identified through daily review of laboratory reports and electronic medical records by the research team to minimize selection bias. Participants were classified into two groups according to the presence of encephalopathic features:

#### VHE group (n = 9)

Patients who developed new acute or subacute disturbance of consciousness (lethargy, stupor, or coma) and/or neuropsychiatric abnormalities (confusion, agitation, or apathy) or cognitive decline during VPA treatment, after exclusion of other potential causes such as intracranial infection, acute cerebrovascular events, electrolyte disturbances, or intracranial space-occupying lesions.

#### Asymptomatic hyperammonemia group (n = 16)

Patients with elevated serum ammonia but without any of the above encephalopathic features.

Exclusion criteria were: (1) severe liver disease (e.g., cirrhosis, acute liver failure) or severe renal disease (chronic kidney disease stages 4–5); (2) concurrent metabolic encephalopathy (e.g., hepatic encephalopathy, uremic encephalopathy); (3) concomitant use of other drugs that may affect ammonia metabolism (e.g., asparaginase, certain antineoplastic agents); and (4) incomplete clinical data.

### 2.3. Data collection and laboratory measurements

Data were extracted from the electronic medical records using a standardized case report form. Collected variables included age; sex (defined as the biological sex assigned at birth and recorded in the medical record); duration of epilepsy; VPA treatment history (total duration of VPA therapy, current daily dose, and recent dose adjustments); concomitant anti-seizure medications; and comorbidities such as hypertension, diabetes mellitus, and cerebrovascular disease.

Serum ammonia was measured using an enzymatic method with glutamate dehydrogenase (institutional reference range, 9–33 μmol/L). Alanine aminotransferase (ALT) and aspartate aminotransferase (AST) were measured using enzymatic rate assays (institutional reference ranges, 7–40 U/L and 13–35 U/L, respectively). VPA trough concentrations were determined by chemiluminescent immunoassay (institutional therapeutic range, 50–100 mg/L). All laboratory assays were performed in the same central laboratory using standardized protocols throughout the study period to ensure comparability between groups.

### 2.4. Electroencephalography

Routine EEG was performed during the symptomatic period using the international 10–20 electrode placement system. EEG findings were classified into three categories based on background rhythm characteristics [16] : (1) normal: posterior dominant α rhythm (8–13 Hz) with well-preserved modulation and amplitude, without excessive slow activity; (2) mild slowing: background rhythm mildly slowed, predominantly increased θ activity, with a still recognizable α rhythm; and (3) diffuse slowing: markedly slowed background rhythm with widespread bilateral θ and/or δ activity and essentially absent α rhythm.

### 2.5. Brain magnetic resonance imaging

Brain magnetic resonance imaging (MRI), including T1-weighted, T2-weighted, and fluid-attenuated inversion recovery (FLAIR) sequences, was performed using standard clinical protocols. Two neuroradiologists who were blinded to group allocation independently reviewed all images, and disagreements were resolved by consensus. The following features were assessed:

Cerebral atrophy (present/absent): defined as reduced brain parenchymal volume, indicated by widening of sulci and/or ventricular enlargement [17];

White matter lesions: graded using the Fazekas scale (0–3); a score of ≥2 was considered moderate-to-severe white matter disease [18]; Old cerebral infarction (present/absent): defined as well-demarcated lesions with low signal on T1WI and high signal on T2WI/FLAIR, without mass effect or enhancement[19].

### 2.6. Treatment and outcome assessment

All patients in the VHE group had VPA discontinued. In the asymptomatic hyperammonemia group, VPA was discontinued, reduced, or maintained at the previous dose based on clinical judgment of the treating physician. No patient received ammonia-lowering therapy (e.g., L-carnitine or lactulose). Serum ammonia was rechecked on days 3, 5, and 7 after the intervention, or before discharge. Symptomatic improvement was defined as recovery of consciousness sufficient to follow simple commands, and complete recovery was defined as return to a normal level of mental status and communication.

### 2.7. Statistical analysis

Statistical analyses were performed using SPSS [version 28.0.1.1, IBM Corp., Armonk, NY, USA]. Because this was an exploratory study with a limited sample size and most continuous variables were not normally distributed (assessed using the Shapiro–Wilk test), nonparametric methods were used throughout. Continuous variables are presented as median (range) and were compared between groups using the Mann–Whitney U test. Categorical variables are presented as counts (percentages) and were compared using Fisher’s exact test.

Between-group comparisons were performed for the following variables: demographics (age, sex); clinical characteristics (duration of epilepsy, VPA treatment history [total duration of VPA therapy, current daily dose, and recent dose adjustment], concomitant anti-seizure medications, and comorbidities); laboratory parameters (serum ammonia, ALT, AST, and VPA trough concentration); EEG background categories; and neuroimaging features (cerebral atrophy, white matter lesion grade, and old cerebral infarction). All tests were two-tailed, and P < 0.05 was considered statistically significant. Given the small sample size and exploratory design, multivariable regression was not performed to avoid overfitting; confounding was addressed through strict exclusion criteria and descriptive stratification. No subgroup or interaction analyses were prespecified. Missing data were minimal due to the requirement of complete clinical records for enrollment; no imputation was applied. Sensitivity analyses were not conducted given the hypothesis-generating nature of this study.

## 3. Results

### 3.1. Participant flow and baseline characteristics

During the study period, 32 consecutive inpatients with epilepsy receiving VPA were screened for eligibility. Of these, 4 were excluded due to severe liver or renal disease, 2 due to concurrent metabolic encephalopathy, and 1 due to incomplete clinical records. The remaining 25 eligible patients were enrolled and included in the final analysis, comprising 9 (36.0%) with VHE and 16 (64.0%) with asymptomatic hyperammonemia. No participants were lost to follow-up or excluded after enrollment. Age, sex, number of comorbidities, VPA daily dose, and VPA trough concentration were comparable between the two groups (all P > 0.05). However, the VHE group had a significantly shorter duration of VPA therapy than the asymptomatic group (median 0 [range 0–60] vs 30 [0–120] months; P = 0.005). ALT and AST levels were within the normal range in both groups, with no between-group differences (P > 0.05) (Table 1). No missing data were observed for any variable of interest among the 25 enrolled participants.

**Table 1.** Baseline and clinical characteristics of the VHE and asymptomatic hyperammonemia (Asym-HA) groups.

| Variable | VHE group<br>(n = 9) | Asym-HA group<br>(n = 16) | P value |
| --- | --- | --- | --- |
| Age, years | 64 (47–80) | 66 (30–82) | 0.777 |
| Male, n (%) | 6 (66.7) | 7 (43.8) | 0.411 |
| Number of comorbidities | 1 (0–3) | 1.5 (0–3) | 0.501 |
| VPA treatment duration, months | 0 (0–60) | 30 (0–120) | 0.005 |
| VPA daily dose, g/day | 1.0 (0.6–1.5) | 1.0 (0.5–1.5) | 0.860 |
| VPA trough concentration, mg/L | 56.7 (39.5–98.7) | 52.2 (37.0–85.7) | 0.412 |
| ALT, U/L | 21.0 (7.0–36.0) | 20.9 (6.2–38.3) | 1.000 |
| AST, U/L | 22.4 (12.6–34.7) | 19.9 (8.0–36.1) | 0.610 |

**Note:** Data are presented as median (range) for continuous variables and n (%) for categorical variables. ALT, alanine aminotransferase; AST, aspartate aminotransferase; Asym-HA, asymptomatic hyperammonemia; VHE, valproate-induced hyperammonemic encephalopathy; VPA, valproic acid. Continuous variables were compared using the Mann-Whitney U test, and categorical variables were compared using Fisher's exact test. A two-tailed $P < 0.05$ was considered statistically significant.

### 3.2. EEG and neuroimaging features

EEG revealed diffuse background slowing in 6 of 9 VHE patients (66.7%) and mild slowing in 3 (33.3%); none had a normal background rhythm. In contrast, 10 asymptomatic patients (62.5%) had a normal EEG, 6 (37.5%) had mild slowing, and none had diffuse slowing (diffuse slowing, 66.7% vs 0%, P < 0.001; normal rhythm, 0% vs 62.5%, P = 0.003).

On brain MRI, the prevalence of cerebral atrophy (44.4% vs 25.0%; P = 0.394) and moderate-to-severe white matter lesions (44.4% vs 50.0%; P = 1.000) did not differ significantly between groups. However, prior cerebral infarction was significantly more common in the VHE group (88.9% vs 43.8%; P = 0.040) (Table 2). These neurophysiological and structural differences prompted us to examine the clinical trajectory of VHE and the outcomes of asymptomatic hyperammonemia.

**Table 2.**
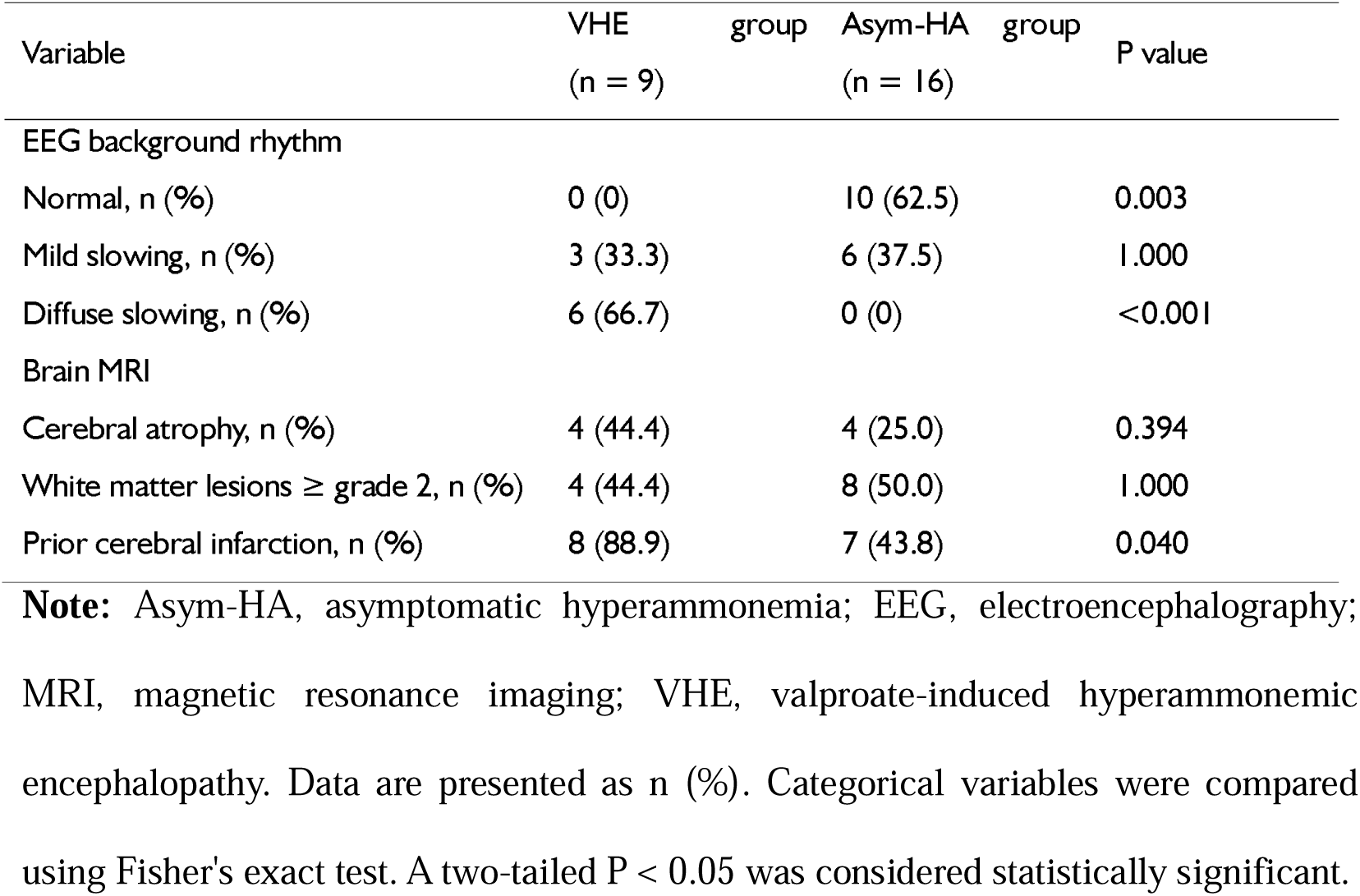
EEG background rhythm and brain MRI characteristics of the VHE and Asym-HA groups.

### 3.3. Clinical course and outcomes of VHE

In the VHE group, encephalopathy developed 2–6 days (median 4 days) after VPA initiation or dose escalation. Six patients (66.7%) developed encephalopathy upon first exposure to VPA; the remaining three had been on long-term VPA (8, 24, and 60 months) and became symptomatic 3–5 days after a recent dose increase. The interval to encephalopathy showed no clear relationship with VPA daily dose (0.6–1.5 g/day) or trough concentration (39.5–98.7 mg/L). Four patients (44.4%) were receiving concomitant anti-seizure medications (levetiracetam in two, perampanel in one, lamotrigine in one) (Table 3).

**Table 3.** Clinical characteristics of the nine patients with VHE.

| Case | VPA duration (mo) | Time to onset (d) | VPA dose (g/d) | Serum VPA (mg/L) | Ammonia (μmol/L) | Concomitant ASM | Symptom improvement (d) | Ammonia normalization (d) |
| --- | --- | --- | --- | --- | --- | --- | --- | --- |
| 1 | 0 | 5 | 0.6 | 56.7 | 82.5 | Lamotrigine | 2 | 5 |
| 2 | 24 | 5 | 1.2 | 39.5 | 76.0 | None | 1 | 3 |
| 3 | 0 | 4 | 1.0 | 72.8 | 149.4 | None | 2 | 7 |
| 4 | 60 | 4 | 1.5 | 39.6 | 52.9 | Levetiracetam | 1 | 3 |
| 5 | 0 | 2 | 1.0 | 98.7 | 155.7 | None | 3 | 10 |
| 6 | 8 | 3 | 1.2 | 49.2 | 61.0 | Perampanel | 2 | 3 |
| 7 | 0 | 6 | 0.6 | 91.1 | 51.2 | None | 1 | 3 |
| 8 | 0 | 3 | 0.6 | 56.1 | 89.1 | None | 2 | 5 |
| 9 | 0 | 3 | 0.6 | 57.0 | 52.3 | Levetiracetam | 1 | 3 |

Note: ASM, anti-seizure medication; VHE, valproate-induced hyperammonemic encephalopathy; VPA, valproic acid. Time to onset refers to the interval from VPA initiation or dose escalation to the appearance of encephalopathic symptoms.

After VPA withdrawal, all nine patients improved within 1–3 days (median 2 days), and serum ammonia normalized or nearly normalized within 3–10 days (median 3 days). None required carnitine, lactulose, or blood purification therapy.

### 3.4. Clinical features and management of asymptomatic hyperammonemia

All 16 asymptomatic patients remained free of encephalopathic symptoms. Their VPA treatment duration was significantly longer than that of the VHE group (median 30 [0–120] vs 0 [0–60] months; P = 0.005). Serum ammonia ranged from 33.5 to 66.0 μmol/L (median 42.5 μmol/L), with no severe elevations.

VPA was discontinued in 2 patients (12.5%), reduced in 10 (62.5%), and left unchanged in 4 (25.0%). Ammonia normalized before discharge in 14 patients (87.5%) within 2–7 days (median 3 days): 12 after dose reduction or discontinuation, and 2 spontaneously without dose adjustment. The remaining 2 patients (12.5%) had only mild ammonia elevation and were receiving low-dose VPA with a favorable benefit–risk profile; their VPA was continued, and although ammonia remained slightly above the upper limit at discharge, neither developed encephalopathic symptoms during follow-up.

### 3.5. Ammonia levels and shared features

Serum ammonia was markedly higher in the VHE group than in the asymptomatic group (76.0 [51.2–155.7] vs 42.5 [33.5–66.0] μmol/L; P < 0.001) (Fig. 1). In both groups, ALT and AST were generally within or near the normal range and did not differ between groups (P = 1.000 and P = 0.610, respectively). VPA trough concentrations were mostly within or below the therapeutic range (50–100 mg/L), with no between-group difference (P = 0.412), indicating that VPA-related hyperammonemia can occur independently of overt liver injury or supratherapeutic drug levels. Among the structural imaging measures, only prior cerebral infarction was associated with VHE (Table 2), suggesting that pre-existing brain damage may confer particular vulnerability to ammonia neurotoxicity.

**Fig. 1.**
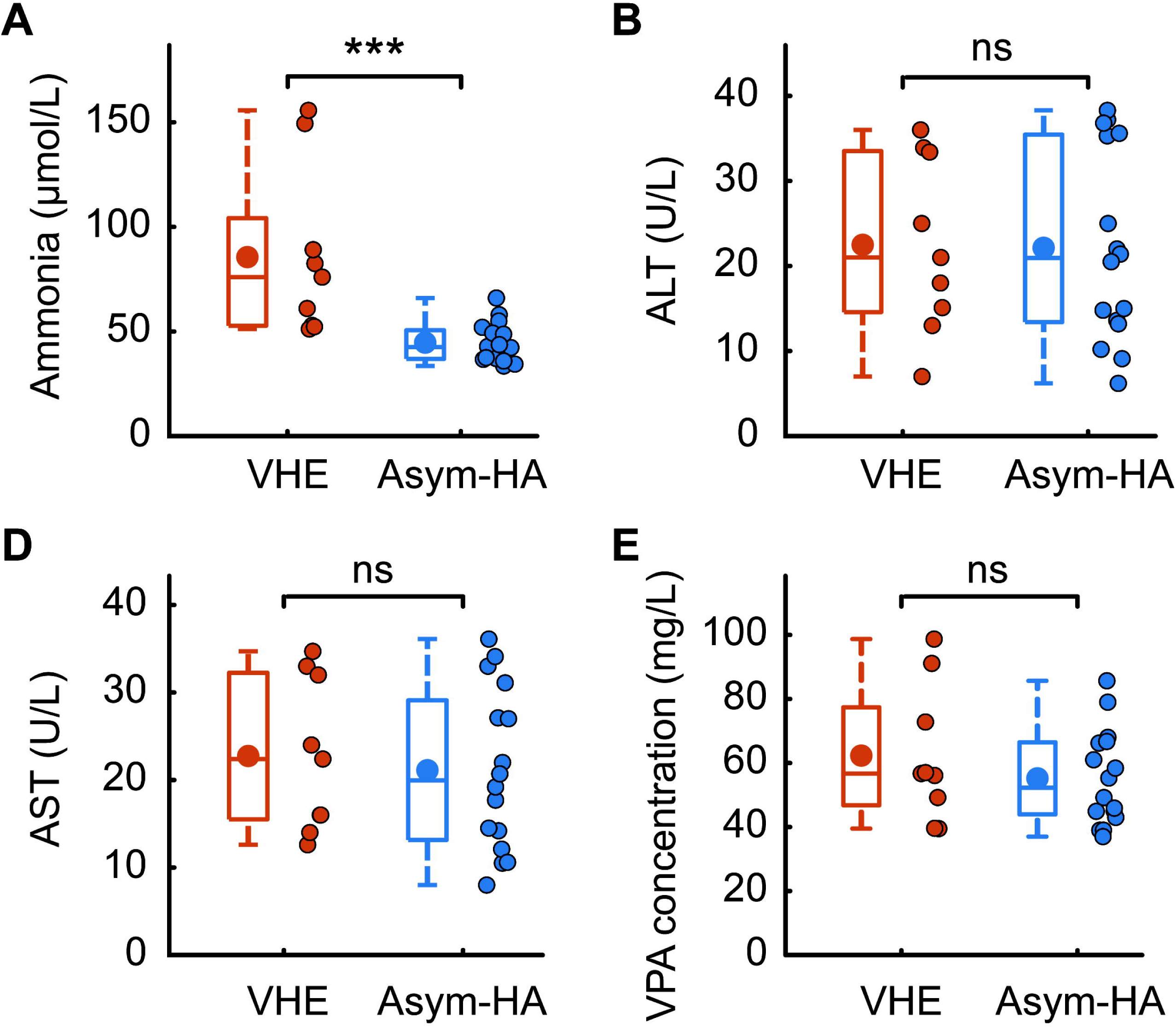
Laboratory parameters in the VHE (n = 9) and Asym-HA (n = 16) groups. **(A)** Serum ammonia; **(B)** alanine aminotransferase (ALT); **(C)** aspartate aminotransferase (AST); **(D)** valproic acid (VPA) concentration. In each panel, the left boxplot shows the median (horizontal line), interquartile range (box), and range (whiskers), with the mean indicated by the central filled circle; jittered dots on the right represent individual patient values. Orange and blue denote the VHE and Asym-HA groups, respectively. P values were calculated using the Mann–Whitney U test: ammonia, P < 0.001; ALT, P = 1.000; AST, P = 0.610; VPA, P = 0.412. Asym-HA, asymptomatic hyperammonemia; VHE, valproate-induced hyperammonemic encephalopathy.

Taken together, VHE was characterized by early onset after VPA initiation or dose escalation, diffuse EEG slowing, a high prevalence of prior cerebral infarction, and rapid recovery after drug withdrawal, whereas asymptomatic hyperammonemia was more common, milder, and usually reversible without VPA discontinuation. No subgroup, interaction, or sensitivity analyses were performed given the exploratory design and limited sample size.

## 4. Discussion

This study of 25 patients with VPA-related hyperammonemia revealed a marked clinical heterogeneity: asymptomatic hyperammonemia accounted for 64.0% (16/25) of cases, outnumbering those with overt VHE (36.0%, 9/25). The VHE group had significantly higher serum ammonia, a shorter duration of VPA therapy, and a higher proportion of diffuse EEG slowing, whereas liver enzymes and VPA trough concentrations did not differ between groups. These findings suggest that VPA-related hyperammonemia spans a spectrum from isolated biochemical abnormality to severe metabolic encephalopathy, requiring stratified approaches to both mechanism and management.

Crucially, our data demonstrate that VHE can occur independently of overt liver injury and supratherapeutic VPA concentrations. Despite comparable ALT, AST, and VPA trough levels (P = 0.412) between groups, serum ammonia was markedly higher in the VHE group. The classic mechanisms of VPA-induced hyperammonemia— inhibition of hepatic mitochondrial carbamoyl phosphate synthetase I, urea cycle disruption, and carnitine depletion with impaired β-oxidation—increase ammonia production and reduce elimination without compromising hepatocellular integrity[20]. Consequently, normal liver enzymes and therapeutic drug levels cannot exclude VHE. From a pharmacological perspective, this underscores a critical limitation of routine therapeutic drug monitoring (TDM): while invaluable for assessing efficacy and avoiding concentration-dependent toxicities, TDM fails to predict metabolically mediated adverse effects such as hyperammonemic encephalopathy. This aligns with previous reports of VHE at therapeutic or subtherapeutic concentrations[21], and warrants a low threshold for ammonia testing in patients developing altered consciousness or neuropsychiatric symptoms during VPA therapy, regardless of liver function or drug levels.

A striking temporal pattern also emerged. The VHE group had a median VPA treatment duration of 0 months, whereas the asymptomatic group had been treated for a substantially longer period. Among the nine VHE patients, six developed encephalopathy upon first exposure to VPA, and three after a recent dose increase following long-term therapy; the interval from initiation or escalation to encephalopathy ranged from 2 to 6 days (median 4 days). This pattern identifies the initiation and dose-titration phases as high-risk periods for VHE. A plausible explanation is that the urea cycle has not yet undergone adaptive regulation during initial therapy or rapid up-titration, so the balance between ammonia production and clearance is rapidly disrupted; in contrast, long-term users may gradually develop metabolic adaptation, manifesting as chronic mild hyperammonemia without progression to encephalopathy. We therefore suggest close monitoring of serum ammonia and level of consciousness during the first week after VPA initiation or dose escalation, and recommend that caregivers be educated about early symptoms such as drowsiness, lethargy, and vomiting.

Notably, prior cerebral infarction was significantly more common in the VHE group than in the asymptomatic group (88.9% vs 43.8%; P = 0.040), whereas cerebral atrophy and white matter lesions were comparable. Moreover, serum ammonia levels overlapped considerably between groups (lowest in VHE: 51.2 μmol/L; highest in asymptomatic: 66.0 μmol/L), indicating that the degree of hyperammonemia is not the sole determinant of encephalopathy. A previous cerebral infarction may cause neuronal loss and disruption of neural circuits, reducing the functional reserve of the central nervous system[22]; at equivalent ammonia levels, individuals with reduced cerebral reserve may be more vulnerable to decompensation. We therefore hypothesize that pre-existing cerebrovascular lesions lower the brain’s tolerance threshold to hyperammonemia—a “reduced cerebral reserve–increased ammonia susceptibility” hypothesis[23, 24] . Given the limited sample size, this association should be regarded as preliminary and requires confirmation in larger prospective studies; nonetheless, heightened vigilance is warranted in VPA users with comorbid cerebrovascular disease.

The EEG findings further support VHE as a diffuse cerebral dysfunction syndrome. Diffuse background slowing was present in 66.7% of VHE patients but in none of the asymptomatic group, and no VHE patient had a normal background rhythm, compared with 62.5% of asymptomatic patients (P = 0.003). Diffuse slowing reflects widespread cortical inhibition and is consistent with the pathophysiology of metabolic encephalopathy[16, 25, 26] . This feature may help differentiate VHE from other causes of altered consciousness such as nonconvulsive status epilepticus or intracranial infection, and may serve as an objective tool for assessing severity and monitoring therapeutic response, particularly in patients who are uncooperative or have atypical presentations.

Our data underscore the cornerstone role of prompt VPA withdrawal in managing VHE. All nine VHE patients improved within 1–3 days (median 2 days) and ammonia normalized or nearly normalized within 3–10 days (median 3 days) after discontinuation, without requiring carnitine, lactulose, or blood purification. This indicates that VHE is largely reversible when recognized early[27].

For asymptomatic hyperammonemia, ammonia normalized before discharge in 14 of 16 patients (87.5%), including two who recovered without any dose adjustment; the remaining two continued VPA because of mild elevation and clear clinical benefit, and neither developed encephalopathic symptoms during follow-up. Thus, asymptomatic hyperammonemia may represent a relatively benign state that does not require aggressive intervention. We recommend immediate VPA discontinuation with supportive care for symptomatic VHE, and a shared decision on dose reduction versus continuation for asymptomatic patients based on the degree of ammonia elevation, therapeutic benefit, and coexisting risk factors, with regular ammonia monitoring. Whether asymptomatic hyperammonemia can progress to encephalopathy in the longer term requires prospective follow-up.

This study has several limitations. First, the single-center design and small sample size (n = 25) preclude multivariable adjustment and increase type II error risk for non-significant findings; however, large effect sizes for key variables (ammonia, EEG slowing, prior infarction) suggest these associations are unlikely to be spurious. Second, the cross-sectional design without long-term follow-up precludes determination of progression risk from asymptomatic hyperammonemia to VHE. Third, retrospective ascertainment of encephalopathy onset may have underestimated the time-to-onset interval if subtle symptoms were undocumented, though this would not affect between-group comparisons. Fourth, fixed-interval ammonia monitoring and non-standardized EEG recording conditions may limit precision in capturing dynamic recovery and background activity comparability, respectively. Fifth, symptom resolution was assessed subjectively. Finally, inclusion of only hospitalized patients may overrepresent severe cases; nonetheless, the consistency of our core findings with existing literature supports their external validity across similar adult epilepsy populations receiving VPA therapy.

In summary, VPA-related hyperammonemia shows considerable clinical heterogeneity: asymptomatic hyperammonemia is common among long-term users, whereas VHE occurs predominantly in patients starting VPA or after recent dose escalation and is characterized by markedly elevated ammonia, diffuse EEG slowing, and a higher prevalence of prior cerebral infarction. VHE can develop independently of liver dysfunction and drug level abnormalities. Clinicians should have a low threshold for ammonia testing and consider VPA withdrawal in patients who develop new-onset alterations in consciousness or behavior during VPA therapy. Future multicenter prospective cohort studies are needed to clarify the natural course of asymptomatic hyperammonemia and to identify predictors of VHE, thereby enabling a more practical risk-stratification and monitoring strategy.

## Data Availability

ll data produced in the present study are available upon reasonable request to the authors

## Declarations

### Availability of data and material

The datasets generated and/or analyzed during the current study are available from the corresponding author upon reasonable request. The data are not publicly available due to privacy and ethical restrictions. Researchers who wish to access the data should contact the corresponding author (X. Wang) for further information.

### Ethics approval

This study was conducted in accordance with the Declaration of Helsinki. The study protocol was approved by the Ethics Committee of the Affiliated Hospital of Jiangsu University (Approval No: KY2024K1102).

### Consent to participate

Written informed consent was obtained from all participants or their legal guardians prior to enrollment.

### Consent for publication

Not applicable.

### Code availability

Not applicable.

### Author contributions

Huijuan Chen and Xuan Wang contributed to the conception and design of the study. Huijuan Chen was responsible for data acquisition, drafting the initial manuscript, and completing the ethics approval application. Xuan Wang performed the data analysis and interpretation, and critically revised the manuscript for important intellectual content. Both authors read and approved the final version of the manuscript and agreed to be accountable for all aspects of the work.

## Acknowledgements

The authors sincerely thank all the participants for their voluntary participation and cooperation. We are also grateful to the clinical and nursing staff of the Departments of Neurology, the Health Examination Center, and Gynecology at the Affiliated Hospital of Jiangsu University for their dedicated assistance in participant recruitment, data collection, and sample processing. Written permission has been obtained from all individuals and entities named in this section.

## References

1. Nieto-Salazar MA, Velasquez-Botero F, Toro-Velandia AC, Saldana-Rodriguez EA, Rodriguez-Rodriguez ME, Gupta A, et al. Diagnostic implications of neuroimaging in epilepsy and other seizure disorders. Ann Med Surg (Lond). 2023 Feb;85(2):73–5.

2. Wang WZ, Wu JZ, Wang DS, Dai XY, Yang B, Wang TP, et al. The prevalence and treatment gap in epilepsy in China: an ILAE/IBE/WHO study. Neurology. 2003 2003/05//;60(9):1544–5.

3. Ding D, Zhou D, Sander JW, Wang W, Li S, Hong Z. Epilepsy in China: major progress in the past two decades. Lancet Neurol. 2021 2021/04//;20(4):316–26.

4. Song P, Liu Y, Yu X, Wu J, Poon AN, Demaio A, et al. Prevalence of epilepsy in China between 1990 and 2015: A systematic review and meta-analysis. J Glob Health. 2017 2017/12//;7(2):020706.

5. Global, regional, and national burden of neurological disorders, 1990-2016: a systematic analysis for the Global Burden of Disease Study 2016. Lancet Neurol. 2019 May;18(5):459–80.

6. Neri S, Gasparini S, Pascarella A, Santangelo D, Cianci V, Mammì A, et al. Epilepsy in Cerebrovascular Diseases: A Narrative Review. Curr Neuropharmacol. 2023;21(8):1634–45.

7. Glauser T, Ben-Menachem E, Bourgeois B, Cnaan A, Guerreiro C, Kälviäinen R, et al. Updated ILAE evidence review of antiepileptic drug efficacy and effectiveness as initial monotherapy for epileptic seizures and syndromes. Epilepsia. 2013 Mar;54(3):551–63.

8. Schiavo A, Maldonado C, Vázquez M, Fagiolino P, Trocóniz IF, Ibarra M. Quantitative systems pharmacology Model to characterize valproic acid-induced hyperammonemia and the effect of L-carnitine supplementation. Eur J Pharm Sci. 2023 Apr 1;183:106399.

9. Chen W, Wang D, Fu W, Wang J, Chen M, Li J, et al. Valproate-induced hyperammonemic encephalopathy: the role of clinical pharmacists in medication safety—a case report. Journal of Medical Case Reports. 2025 2025/07/01;19(1):302.

10. Wu MY, Chang FY, Ke JY, Chen CS, Lin PC, Wang TS. Valproic Acid-Induced Hyperammonemic Encephalopathy in a Patient with Bipolar Disorder: A Case Report. Brain Sci. 2020 Mar 24;10(3).

11. Aires C, van Cruchten AG, Ijlst L, de Almeida IT, Durán M, Wanders RJA, et al. New insights on the mechanisms of valproate-induced hyperammonemia: inhibition of hepatic N-acetylglutamate synthase activity by valproyl-CoA. Journal of hepatology. 2011;55 2:426–34.

12. Chopra A, Kolla BP, Mansukhani MP, Netzel PJ, Frye MA. Valproate-induced hyperammonemic encephalopathy: an update on risk factors, clinical correlates and management. General hospital psychiatry. 2012;34 3:290–8.

13. Segura-Bruna N, Rodriguez-Campello A, Puente V, Roquer J. Valproate-induced hyperammonemic encephalopathy. Acta Neurol Scand. 2006 Jul;114(1):1–7.

14. Wadzinski J, Franks R, Roane D, Bayard M. Valproate-associated Hyperammonemic Encephalopathy. The Journal of the American Board of Family Medicine. 2007;20(5):499–502.

15. Carr RB, Shrewsbury K. Hyperammonemia due to valproic acid in the psychiatric setting. Am J Psychiatry. 2007 2007/07//;164(7):1020-7.

16. Hirsch LJ, Fong MWK, Leitinger M, LaRoche SM, Beniczky S, Abend NS, et al. American Clinical Neurophysiology Society’s Standardized Critical Care EEG Terminology: 2021 Version. J Clin Neurophysiol. 2021 Jan 1;38(1):1–29.

17. Lim KY, Park S, Na DL, Seo SW, Chun MY, Kwak K. Quantifying Brain Atrophy Using a CSF-Focused Segmentation Approach. Dement Neurocogn Disord. 2025 Apr;24(2):115–25.

18. Xing Y, Yang J, Zhou A, Wang F, Tang Y, Jia J. Altered brain activity mediates the relationship between white matter hyperintensity severity and cognition in older adults. Brain Imaging Behav. 2022 Apr;16(2):899–908.

19. Zhang L, Yu X, Zheng Y, Lin A, Zhang Z, Li S, et al. Lacunes are associated with late-stage multiple sclerosis comorbidities. Front Neurol. 2023;14:1224748.

20. Wu J, Li J, Jing W, Tian X, Wang X. Valproic acid-induced encephalopathy: A review of clinical features, risk factors, diagnosis, and treatment. Epilepsy Behav. 2021 Jul;120:107967.

21. Sammar A, Tawfik M, Fatima F, Butler A, Aylor-Lee K. Valproate-Induced Hyperammonemic Encephalopathy Causing New-Onset Seizures. Cureus. 2023 Oct;15(10):e47288.

22. Brown GC. Neuronal Loss after Stroke Due to Microglial Phagocytosis of Stressed Neurons. Int J Mol Sci. 2021 Dec 14;22(24).

23. Cordova-Gallardo J, Vargas-Beltran AM, Armendariz-Pineda SM, Ruiz-Manriquez J, Ampuero J, Torre A. Brain reserve in hepatic encephalopathy: Pathways of damage and preventive strategies through lifestyle and therapeutic interventions. Ann Hepatol. 2025 Jan-Jun;30(1):101740.

24. Jokinen H, Melkas S, Madureira S, Verdelho A, Ferro JM, Fazekas F, et al. Cognitive reserve moderates long-term cognitive and functional outcome in cerebral small vessel disease. J Neurol Neurosurg Psychiatry. 2016 2016/12//;87(12):1296–302.

25. Yýlmaz A, Yayıcı Köken Ö, Şekeroğlu B, Şanlıdağ B. A Near-Global Slowing of Background Activity and Epileptic Discharges in Children With Mild to Moderately Symptomatic COVID-19 Infection: An Electro-Neurophysiological Study. Clin EEG Neurosci. 2022 Nov;53(6):532–42.

26. Kaplan PW. The EEG in metabolic encephalopathy and coma. Journal of clinical neurophysiology : official publication of the American Electroencephalographic Society. 2004 2004 Sep-Oct;21(5):307–18.

27. Liu X, Peng X. Valproate-related hyperammonemic encephalopathy with generalized suppression EEG: a case report. Neurol Sci. 2023 Oct;44(10):3669–73.

